# Visual-Textual Integration in LLMs for Medical Diagnosis: A Quantitative Analysis

**DOI:** 10.1101/2024.08.31.24312878

**Authors:** Reem Agbareia, Mahmud Omar, Shelly Soffer, Benjamin S Glicksberg, Girish N Nadkarni, Eyal Klang

## Abstract

**Background and Aim:** Visual data from images is essential for many medical diagnoses. This study evaluates the performance of multimodal Large Language Models (LLMs) in integrating textual and visual information for diagnostic purposes.

**Methods:** We tested GPT-4o and Claude Sonnet 3.5 on 120 clinical vignettes with and without accompanying images. Each vignette included patient demographics, a chief complaint, and relevant medical history. Vignettes were paired with either clinical or radiological images from two sources: 100 images from the OPENi database and 20 images from recent NEJM challenges, ensuring they were not in the LLMs’ training sets. Three primary care physicians served as a human benchmark. We analyzed diagnostic accuracy and the models’ explanations for a subset of cases.

**Results:** LLMs outperformed physicians in text-only scenarios (GPT-4o: 70.8%, Claude Sonnet 3.5: 59.5%, Physicians: 39.5%). With image integration, all improved, but physicians showed the largest gain (GPT-4o: 84.5%, p<0.001; Claude Sonnet 3.5: 67.3%, p=0.060; Physicians: 78.8%, p<0.001). LLMs changed their explanations in 45-60% of cases when presented with images, demonstrating some level of visual data integration.

**Conclusion:** Multimodal LLMs show promise in medical diagnosis, with improved performance when integrating visual evidence. However, this improvement is inconsistent and smaller compared to physicians, indicating a need for enhanced visual data processing in these models.

## Introduction

Visual data, such as direct patient examination or medical imaging, is vital for patient diagnostics (1). Multimodal capabilities in LLMs, introduced around 2021, allow these models to process both text and visual inputs (2,3). This is achieved through techniques like vision transformers and cross-attention mechanisms, which enable the model to align visual features with textual information (4). For example, a multimodal LLM can analyze both a patient’s written symptoms and an X-ray image, potentially enhancing diagnostic accuracy (5).

While the integration of visual information into LLMs holds promise, its application in healthcare remains understudied. It is unclear how effectively these models incorporate visual clinical and imaging data alongside textual information when making medical diagnoses (6,7).

A key question is whether LLMs tend to prioritize textual information over visual cues (8). Alternatively, they might equally weigh both modalities, or potentially underutilize visual data. Understanding this balance is important for optimizing the use of multimodal LLMs in medical applications.

This study evaluates the performance of multimodal LLMs in integrating textual and visual information for diagnostic purposes.

## Materials and Methods

### Study Design and Data Preparation

We evaluated LLMs’ performance in diagnosing clinical cases using textual and visual data. Our dataset comprised 120 clinical vignettes, each with patient demographics, chief complaint, and relevant history. We included 100 images (80 clinical and 20 radiological images) from OPENi (https://openi.nlm.nih.gov). We also included 20 images from NEJM challenges published after March 2024, ensuring that they were not part of the LLMs’ training data (**Figure 1** presents an example case).

**Figure 1:**
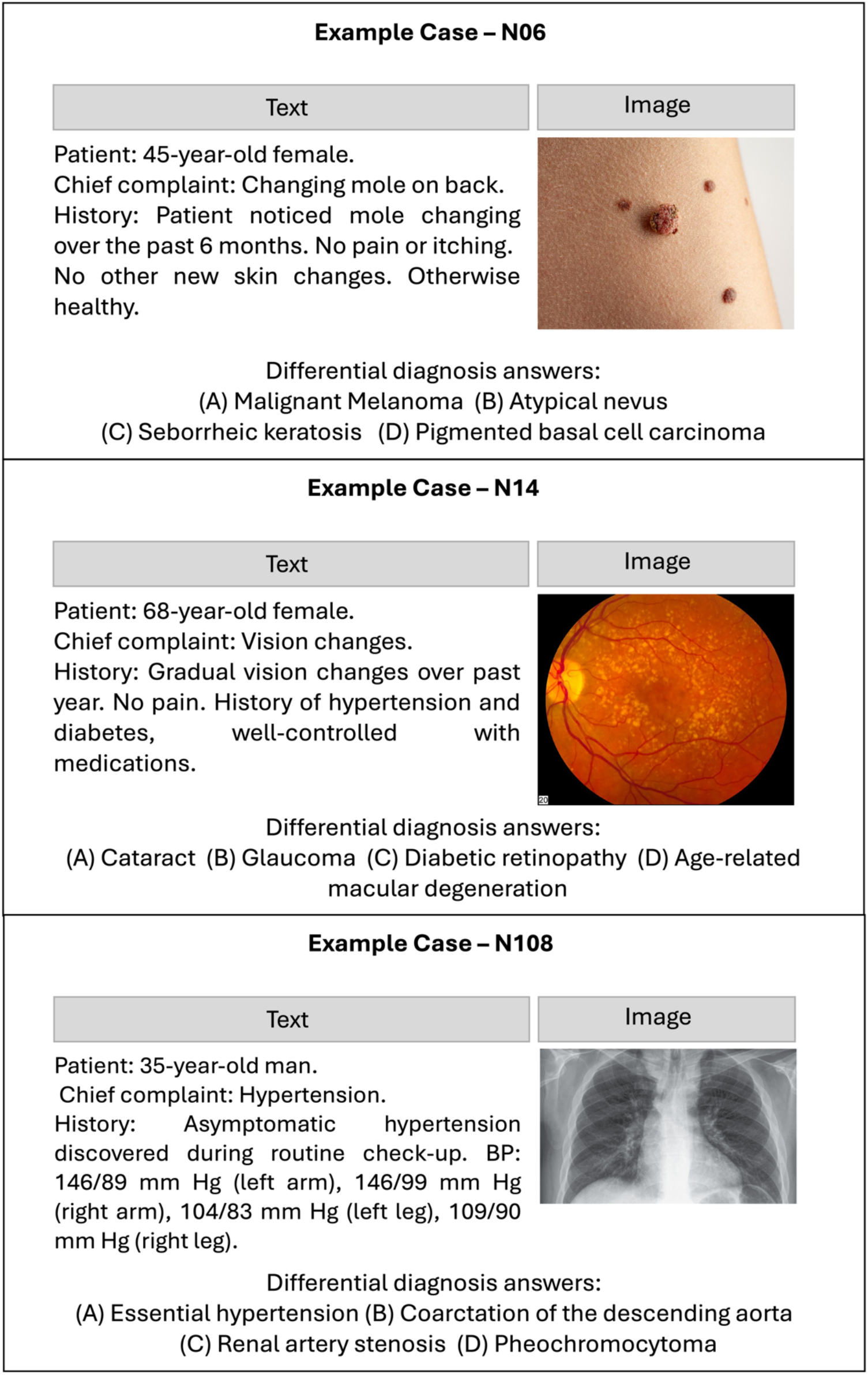
Example cases.

Each case was built around a pre-selected image, encompassing various medical fields and imaging modalities. We used a standardized, short form for the vignettes, avoiding evident textual data that could lead directly to diagnosis without image integration. Four differential diagnosis options were carefully chosen for each case. Two doctors wrote and cross-validated each case, following epidemiological guides for chief complaints and presenting symptoms (9). We adhered to a systematic and consistent guidelines for creating each clinical vignettes (10), and used GPT-4 API for proofreading and validation (11). (A complete flowchart of creating the case vignettes can be found in the **supplement**).

For a subset of 40 cases, we asked the models to provide explanations for their diagnoses in both text-only and text + imaging scenarios. We defined a “changed explanation” as when the model provides significantly different reasoning or details in its explanation for the text + imaging scenario compared to the text-only scenario, regardless of whether the final diagnosis changed. Two physicians evaluated these explanations, assessing whether the model mentioned using visual data and comparing the explanations between scenarios.

### Model Selection and Implementation

We selected three multimodal LLMs for our study: GPT-4o and Claude Sonnet 3.5. The implementation was carried out using Python 3.9, utilizing the OpenAI API (version 1.3.5) for GPT models and the Anthropic API (version 0.2.8) for Claude Sonnet 3.5. Data processing and numerical operations were performed using Pandas (version 1.5.3) and NumPy (version 1.23.5) libraries.

Each case was presented to the models twice: with text only and with text and image combined. We used a standardized prompt across all models, found in the **Supplement**. For image inputs, we employed base64 encoding to include images in the API calls.

### Human Benchmark

To benchmark against human performance, three board-certified primary care physicians independently evaluated all cases in both the text-only and the text + imaging formats. Their responses were collected using a custom-built web interface to ensure consistency in presentation.

### Data Analysis

We calculated descriptive statistics, including the number of correct answers for each model and human physicians, separately for text-only and text + imaging inputs. We performed paired t-tests to compare performance between text-only and text + imaging conditions for each model and human physicians. One-way ANOVA was used to compare the difference in performance improvement across all models and human physicians, followed by post-hoc pairwise t-tests with Bonferroni correction for multiple comparisons.

We calculated the magnitude of difference between text-only and text + imaging conditions by comparing the number of correct answers in each condition. Subgroup analysis and Cohen’s kappa coefficient calculations were not performed in this study. The R packages used for analysis included tidyverse (version 1.3.2) and stats (version 4.2.2). All statistical tests were two-tailed with a significance level of α = 0.05, and Bonferroni correction was applied for multiple comparisons where necessary.

## Results

### Model and Physicians Overall Performance

Across the combined datasets, GPT-4o’s overall performance improved from 70.8% (±12.95%) to 84.5% (±7.75%) with image integration, a 13.7% increase (p < 0.001). Claude Sonnet 3.5 improved from 59.5% (±12.95%) to 67.3% (±7.75%), a 7.8% increase (p = 0.06). Physicians showed the largest improvement, from 39.5% (±12.95%) to 78.8% (±7.75%), a 39.3% increase (p < 0.001). In the Full dataset, GPT-4o achieved 75% (±10.6%) without images and 89% (±4.0%) with images, Claude Sonnet 3.5 improved from 64% (±10.6%) to 77% (±4.0%), and physicians from 42.3% (±10.6%) to 70.7% (±4.0%) (**Figure 1**).

**Figure 1:**
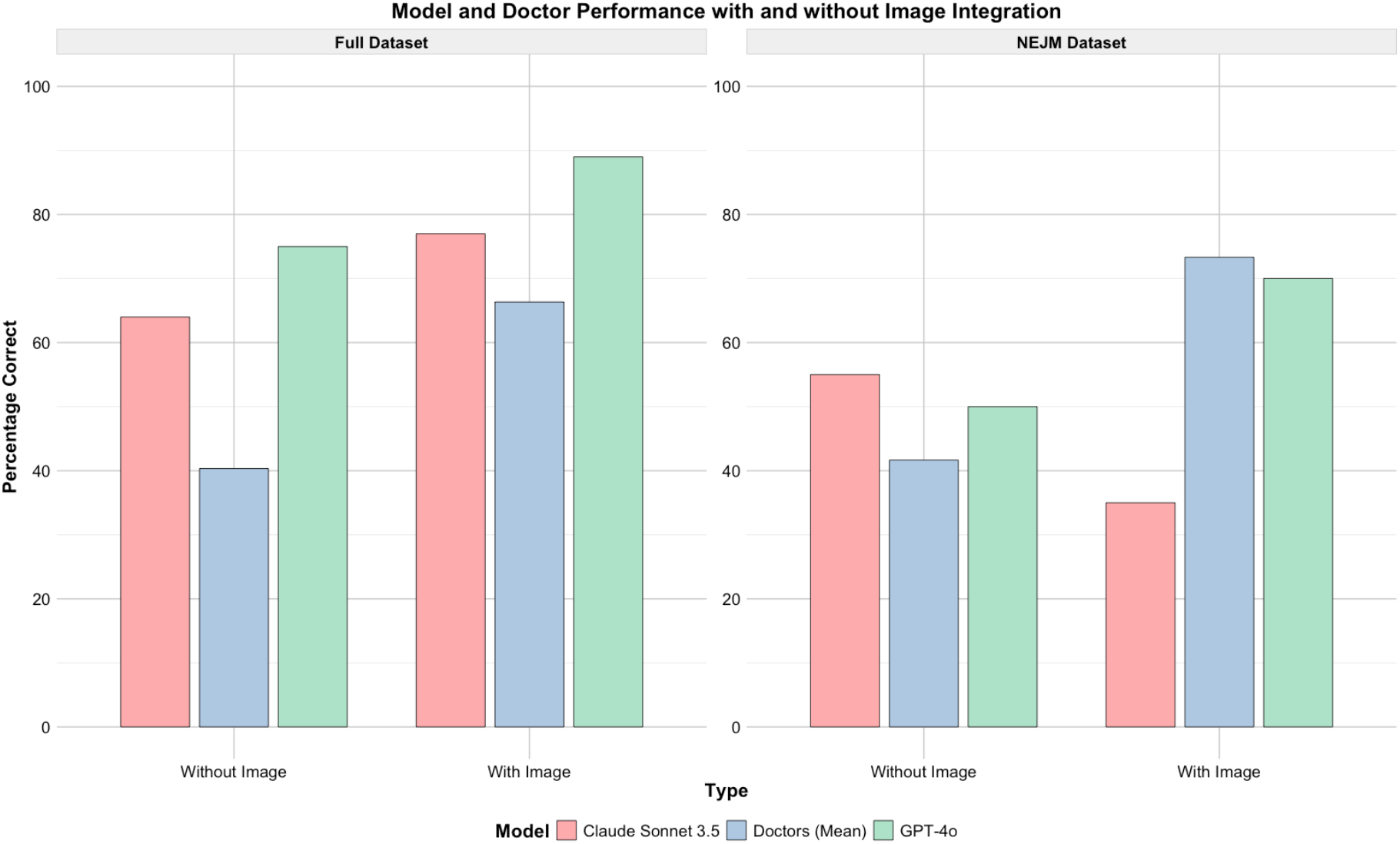
Performances across datasets, models and physicians.

### Performance Change with Image Integration

For GPT-4o, performance significantly improved with image integration. In the Full dataset, the correct response rate increased by 14% (p < 0.001), and in the NEJM dataset, it increased by 20% (p < 0.001). Claude Sonnet 3.5 showed a 13% improvement in the Full dataset, though this change was not statistically significant (p = 0.060). However, Claude Sonnet 3.5’s performance in the NEJM dataset dropped significantly with image integration, decreasing from 55% correct in the text-alone condition to 35% correct in the text + image condition (p < 0.001).

Across the combined datasets, GPT-4o’s overall performance improved from 70.8% to 84.5%, while Claude Sonnet 3.5 improved from 59.5% to 67.3% (**Table 1**).

**Table 1:**
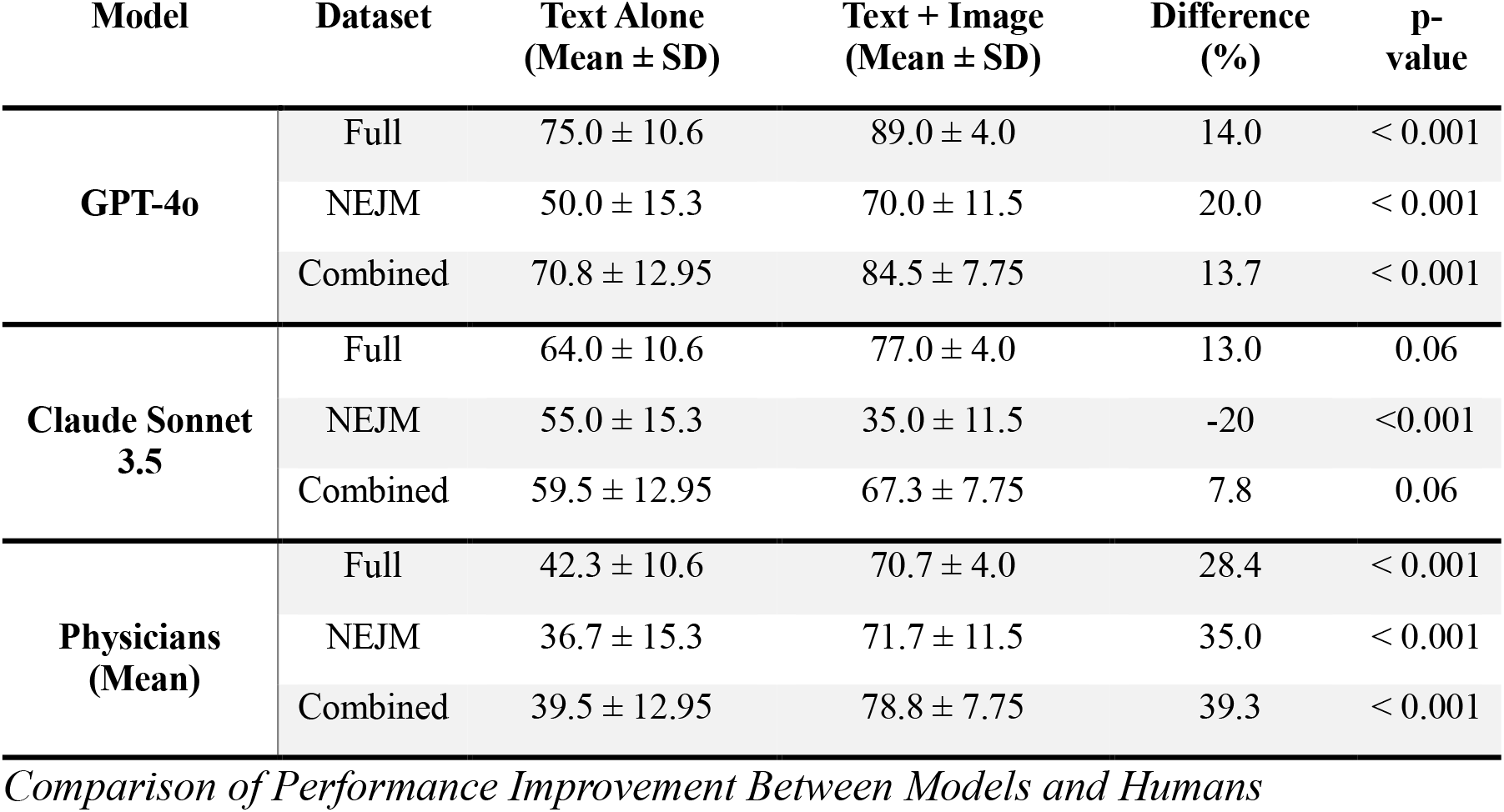
Performance Comparison of Models and Physicians in Text Alone vs. Text + Image Conditions Across Datasets.

### Comparison of Performance Improvement Between Models and Humans

The average improvement in performance for physicians with image integration was 28.3% in the Full dataset and 35% in the NEJM dataset. These improvements were statistically significant (p < 0.001).

In the combined datasets, physicians improved from 39.5% to 78.8%, a 39.3% increase. The improvement for physicians was significantly greater than for GPT-4o in both datasets (p < 0.001), and significantly greater than for Claude Sonnet 3.5 in the NEJM dataset (p < 0.001). There was no significant difference in performance improvement between GPT-4o and Claude Sonnet 3.5 (p = 0.393).

### Analysis of Reasoning

In our analysis of model explanations, GPT-4o changed its explanations in 45% of cases and its diagnosis in 20% of cases when presented with images. Claude Sonnet 3.5 changed its explanations in 60% of cases and diagnoses in 15% of cases. Both models frequently referenced image evidence in their explanations, with GPT-4o doing so in 82.5% of cases and Claude Sonnet 3.5 in 77.5% of cases. The changes in explanations often involved shifts in reasoning, such as GPT-4o changing its diagnosis from malignant melanoma to seborrheic keratosis in a skin lesion case, or Claude Sonnet 3.5 altering its assessment from tension headaches to a brain tumor when presented with an MRI image (**Table 2**).

**Table 2:**
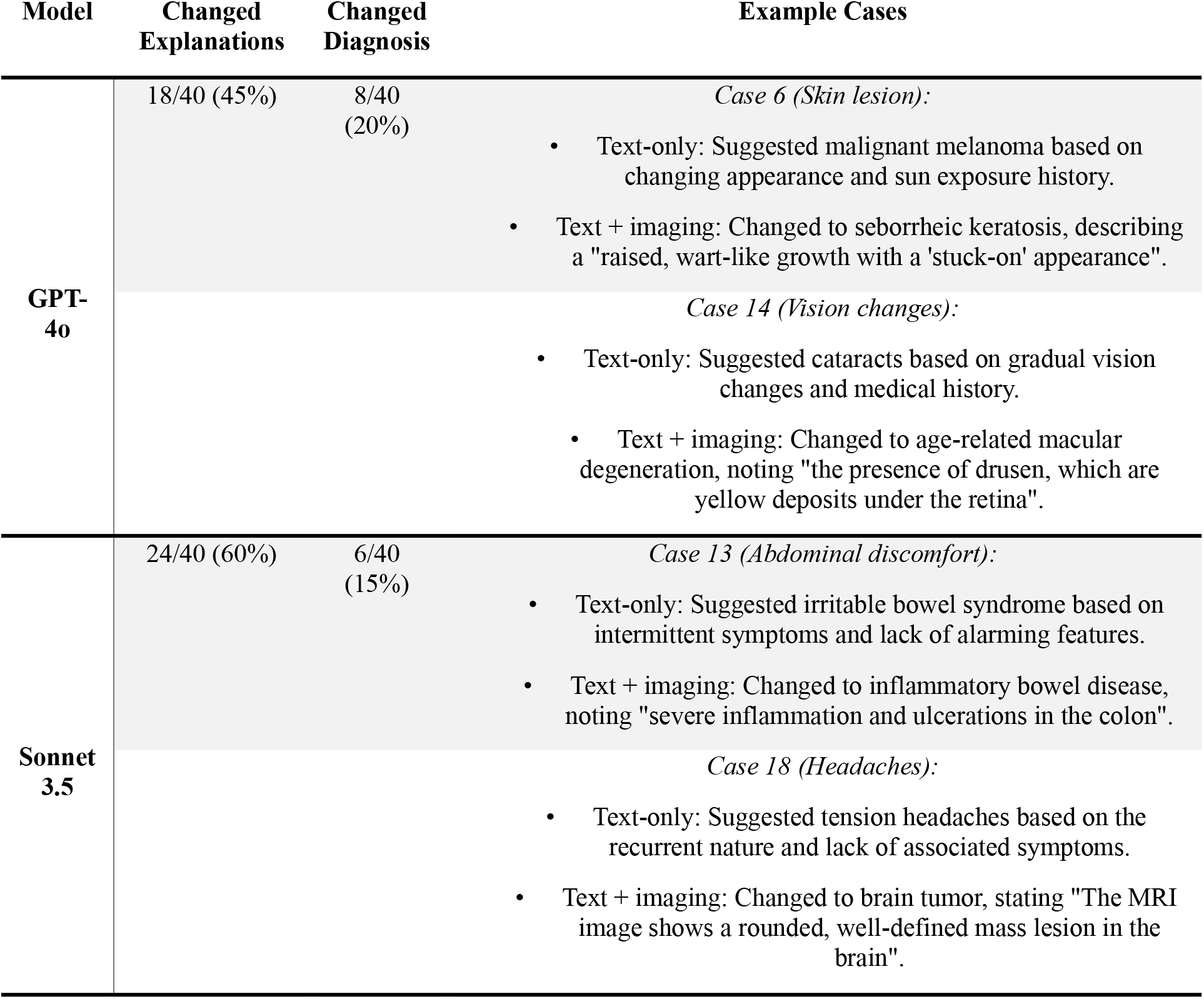
Specific examples of changed explanations between textual only and textual with vision inputs.

## Discussion

Our study reveals insights into the performance of multimodal LLMs in medical diagnosis. Both GPT-4o and Claude Sonnet 3.5 improved diagnostic accuracy when integrating visual data, with GPT-4o showing consistent improvement across datasets, while Claude Sonnet 3.5’s performance varied.

GPT-4o demonstrated improved performance with image integration, suggesting effective synthesis of visual and textual information. In contrast, Claude Sonnet 3.5’s performance declined in the NEJM dataset with images, indicating potential challenges in complex medical imaging interpretation. Analysis of model explanations revealed distinct approaches: GPT-4o maintained consistent detail across modalities, while Claude Sonnet 3.5 provided more extensive explanations with images. These differences highlight varying strategies in visual data integration and reasoning processes between the models.

Notably, LLMs outperformed physicians in text-only scenarios, despite cases being designed for textual and vision diagnosis. This suggests LLMs possess extensive medical knowledge and potentially employ statistical approaches, choosing the most probable answer based on similar textual data in their training (12). This statistical approach might explain some mistakes made even with image integration and the smaller performance leap compared to humans when images were added.

The smaller performance leap of LLMs compared to human physicians when integrating images warrants examination. This disparity might indicate that LLMs use image data less effectively than human experts. Our data shows instances where all three physicians correctly diagnosed cases using images, while one or more LLMs failed. This suggests room for improvement in LLMs’ visual data processing and integration.

The diverse range of medical fields represented in our image dataset, contrasted with our primary care physician panel, presents both a limitation and an insight. While primary care physicians are trained across multiple specialties (13,14), their performance might not fully represent specialist-level image interpretation. However, this scenario mirrors real-world primary care, where generalists encounter a wide range of conditions (13). The LLMs’ performance across these diverse cases showcases their potential as versatile diagnostic support tools across medical specialties.

LLMs outperforming physicians in question answering is well-documented. This aligns with existing research and benchmarks (15–17). However, evidence for LLMs’ multimodal diagnostic abilities is scarce. Some studies, particularly in radiology (18), suggest LLMs perform well in multimodal diagnosis (2,18), yet we found no direct comparisons between multimodal, diverse diagnostic scenarios and text-only performance.

Privacy concerns in medical imaging pose a significant challenge for LLM development. Training these models on large-scale human medical vision data while maintaining patient privacy is complex (19). This limitation highlights the need for innovative approaches to data anonymization and synthetic data generation in medical AI research.

Our study has limitations. The sample size, particularly for the NEJM dataset is small, as it included only recently published cases to ensure that they were not part of the LLMs’ training data. Additionally, artificially constructed cases may not fully represent real-world clinical complexity. Future studies should consider larger, diverse datasets and real-world cases. Moreover, we did not address the analysis of visual data over time, a crucial aspect in many clinical scenarios. The progression of skin rashes or growth of pulmonary nodules, for instance, often guides diagnosis and management. Future research should explore LLMs’ capability to integrate and interpret sequential medical imaging, potentially enhancing their clinical utility.

In conclusion,

Multimodal LLMs show promise in medical diagnosis, with improved performance when integrating visual evidence. However, this improvement is inconsistent and smaller compared to physicians, indicating a need for enhanced visual data processing in these models.

## Data Availability

All data produced in the present study are available upon reasonable request to the authors

## Acknowledgment

We thank the primary care physicians who contributed their time and expertise to benchmark the clinical cases, providing an essential human performance baseline for our study.

