## Supplement material for "Visual-Textual Integration in LLMs for Medical Diagnosis: A Quantitative Analysis"

### Detailed Process of Case Building

We developed a rigorous process for creating and validating our clinical vignettes, as illustrated in Figure S1. This process involved the following steps:

a) Image Selection:

- We selected 100 images from the OPENi database (https://openi.nlm.nih.gov) and 20 images from NEJM challenges published after March 2024.
- Images were chosen to represent various medical fields (e.g., internal medicine, radiology, dermatology) and different types of clinical and radiological modalities.
- We ensured a diverse range of conditions and presentations to test the models' capabilities across various medical scenarios.

b) Vignette Creation:

- Two board-certified doctors independently wrote clinical vignettes for each selected image.
- We used a standardized, short form including demographic data (age and sex), chief complaint, and relevant history.
- Care was taken to avoid including evident textual data that could lead directly to the diagnosis without image integration.

c) Differential Diagnosis:

- For each case, we carefully selected four differential diagnosis options, including the correct diagnosis.
- These options were chosen to be plausible alternatives based on the presented information.

d) Cross-validation:

- The two doctors cross-validated each other's vignettes, ensuring clinical accuracy and adherence to the study guidelines.

e) Epidemiological Alignment:

- We aligned our cases with the epidemiology of chief complaints and presenting symptoms according to Hooker et al. (2019) to ensure realistic representation of clinical scenarios.

f) Vignette Quality Assurance:

- We followed the systematic process outlined by Stacey et al. (2014) for creating and appraising clinical vignettes.

g) AI-assisted Proofreading and Validation:

- We utilized GPT-4 API as a confirming, proofreading, and validating tool for the vignettes, as suggested by Coşkun et al. (2024).

h) Final Review:

- A third doctor reviewed all cases to ensure consistency, clarity, and adherence to study objectives.

### Prompts Used

a) Without Explanation Prompt:

Please think deeply about the following question, integrating all of the data in it, and return the answer. Return your answer in a JSON format with two keys: "case_number" and "diagnosis". The "diagnosis" should be only the letter A, B, C, or D.

Case: {case}

Question: {question}

Answer (in JSON format):

b) With Explanation Prompt:

Please think deeply about the following question, integrating all of the data in it, and return the answer. Return your answer in a JSON format with three keys: "case_number", "diagnosis", and "explanation". The "diagnosis" should be only the letter A, B, C, or D.

The "explanation" should provide a brief rationale for your diagnosis.

Case: {case}

Question: {question}

Answer (in JSON format):

c) System message:

{"role": "system", "content": "You are an expert medical professional. Provide the answer in the requested format without any additional text."}

d) User Message for Text-only Response:

{"role": "user", "content": "What is the correct diagnosis?"}

#### Technical Implementation Details

a) API Calls:

- We used OpenAI API (version 1.3.5) for GPT models and Anthropic API (version 0.2.8) for Claude Sonnet 3.5.
- Python 3.9 was used for implementation, with Pandas (version 1.5.3) and NumPy (version 1.23.5) for data processing.

b) Image Encoding:

- For image inputs, we used base64 encoding to include images in the API calls.

c) Response Processing:

- Responses were parsed from JSON format and stored for analysis.
- We implemented error handling and retries (max 3 attempts) for failed API calls.

d) Data Analysis:

- R (version 4.2.2) with tidyverse (version 1.3.2) and stats (version 4.2.2) packages were used for statistical analysis.
- We performed paired t-tests, one-way ANOVA, and post-hoc pairwise t-tests with Bonferroni correction.

e) Explanation Analysis:

- Two doctors independently reviewed the explanations provided by the models for a subset of 40 cases.
- They assessed whether the model mentioned using visual data and compared explanations between text-only and text+image scenarios.
